# Covid-19 vaccination and menstrual cycle length in the Apple Women’s Health Study

**DOI:** 10.1101/2022.07.07.22277371

**Authors:** Elizabeth A. Gibson, Huichu Li, Victoria Fruh, Malaika Gabra, Gowtham Asokan, Anne Marie Z. Jukic, Donna D. Baird, Christine L Curry, Tyler Fischer-Colbrie, Jukka-Pekka Onnela, Michelle A. Williams, Russ Hauser, Brent A. Coull, Shruthi Mahalingaiah

## Abstract

**Background:** COVID-19 vaccination may be associated with change in menstrual cycle length following vaccination.

**Methods:** We conducted a longitudinal analysis within a subgroup of 14,915 participants in the Apple Women’s Health Study (AWHS) who enrolled between November 2019 and December 2021 and met the following eligibility criteria: were living in the U.S., met minimum age requirements for consent, were English speaking, actively tracked their menstrual cycles, and responded to the COVID-19 Vaccine Update survey. In the main analysis, we included tracked cycles recorded when premenopausal participants were not pregnant, lactating, or using hormonal contraceptives. We used conditional linear regression and multivariable linear mixed-effects models with random intercepts to estimate the covariate-adjusted difference in mean cycle length, measured in days, between pre-vaccination cycles, cycles in which a vaccine was administered, and post-vaccination cycles within vaccinated participants, and between vaccinated and unvaccinated participants. We further compared associations between vaccination and menstrual cycle length by the timing of vaccine dose within a menstrual cycle (i.e., in follicular or luteal phase). We present Bonferroni-adjusted 95% confidence intervals to account for multiple comparisons.

**Results:** A total of 128,094 cycles (median = 10 cycles per participant; interquartile range: 4-22) from 9,652 participants (8,486 vaccinated; 1,166 unvaccinated) were included. The average within-individual standard deviation in cycle length was 4.2 days. Fifty-five percent of vaccinated participants received Pfizer-BioNTech’s mRNA vaccine, 37% received Moderna’s mRNA vaccine, and 7% received the Johnson & Johnson/Janssen vaccine (J&J). We found no evidence of a difference between mean menstrual cycle length in the unvaccinated and vaccinated participants prior to vaccination (0.24 days, 95% CI: −0.34, 0.82).

Among vaccinated participants, COVID-19 vaccination was associated with a small increase in mean cycle length (MCL) for cycles in which participants received the first dose (0.50 days, 95% CI: 0.22, 0.78) and cycles in which participants received the second dose (0.39 days, 95% CI: 0.11, 0.67) of mRNA vaccines compared with pre-vaccination cycles. Cycles in which the single dose of J&J was administered were, on average, 1.26 days longer (95% CI: 0.45, 2.07) than pre-vaccination cycles. Post-vaccination cycles returned to average pre-vaccination length. Estimates for pre vs post cycle lengths were 0.14 days (95% CI: −0.13, 0.40) in the first cycle following vaccination, 0.13 days (95% CI: −0.14, 0.40) in the second, −0.17 days (95% CI: −0.43, 0.10) in the third, and −0.25 days (95% CI: −0.52, 0.01) in the fourth cycle post-vaccination. Follicular phase vaccination was associated with an increase in MCL in cycles in which participants received the first dose (0.97 days, 95% CI: 0.53, 1.42) or the second dose (1.43 days, 95% CI: 1.06, 1.80) of mRNA vaccines or the J&J dose (2.27 days, 95% CI: 1.04, 3.50), compared with pre-vaccination cycles.

**Conclusions:** COVID-19 vaccination was associated with an immediate short-term increase in menstrual cycle length overall, which appeared to be driven by doses received in the follicular phase. However, the magnitude of this increase was small and diminished in each cycle following vaccination. No association with cycle length persisted over time. The magnitude of change associated with vaccination was well within the natural variability in the study population. Menstrual cycle change following COVID-19 vaccination appears small and temporary and should not discourage individuals from becoming vaccinated.

## Introduction

Though the United States Vaccine Adverse Event Reporting System (VAERS) does not actively collect information regarding menstrual cycles, over 11,000 people self-reported a menstrual-related issue to the system following COVID-19 vaccination by April 2022.^1^ Events included heavy bleeding, irregular or delayed menstruation, oligomenorrhea, and amenorrhea. Likewise, the United Kingdom’s Medicines and Healthcare Products Regulatory Agency (MHRA) yellow card surveillance scheme received over 50,000 reports of menstrual disorders, including heavier than usual periods, delayed periods, and unexpected bleeding, after the COVID-19 vaccine.^2^ This anecdotal evidence suggested the possibility of temporary changes in menstrual cycles following COVID-19 vaccination.^3, 4^ Women reported these menstrual changes to their doctors and posted about them on social media.^5, 6^ Various news outlets publicized the anecdotal reports.^7–9^

COVID-19 vaccination is widely regarded as safe,^10, 11^ and there is no evidence that it affects fertility in laboratory models,^12, 13^ clinical trials,^14, 15^ or observational studies.^16–18^ Still, the immune response or the associated stress may link the vaccine with certain temporary menstrual changes.^3^ Improved understanding of these associations, such as whether one exists and the magnitude and persistence of potential changes, will assist clinicians in counselling women concerning vaccination.^19^

While research on the COVID-19 vaccine and menstrual characteristics is limited, existing studies have reported temporarily increased menstrual cycle length,^20^ increased menstrual bleeding,^21^ longer duration of menses,^22^ and menstrual irregularities,^23, 24^ as well as breakthrough bleeding in individuals who previously menstruated.^25^ Generally, these changes were considered short term, but detailed follow-up data beyond the first cycle after vaccination were not presented. These reports suggest the possibility of temporary changes in menstrual cycles following COVID-19 vaccination.^3, 4^

Here, we evaluate the relationship between COVID-19 vaccination and menstrual cycle length over time in the Apple Women’s Health Study (AWHS), a longitudinal digital cohort of people in the U.S. with manually tracked menstrual cycles.^26^ We compare pre-vaccination cycle lengths with those in which a vaccine dose was administered and cycles following vaccination.

## Methods

### Study design and population

The AWHS is a prospective digital cohort of participants in the U.S.. Recruitment began in November 2019 and is currently on-going.^26^ Enrollment eligibility specifies that participants must be assigned female at birth, have menstruated at least once, live in the U.S., be at least 18 years old (at least 19 years old in Alabama and Nebraska and at least 21 years old in Puerto Rico), be able to communicate through written and spoken English, have an iPhone with a compatible version of iOS, download the Apple Research app, be the sole user of an iPhone and an iCloud account, and provide informed consent for participation. This study was approved by the Institutional Review Board at Advarra (CIRB #PRO00037562) and registered on Clinicaltrials.gov (NCT04196595).

Detailed information on study design, participant recruitment, and data collection has been described previously.^26^ Briefly, participants completed surveys on demographic characteristics and baseline menstrual status at enrollment, on reproductive history within the first year, and on health behaviors and gynecological history once a year during follow-up. After enrollment, participants were surveyed monthly regarding their menstrual health. A COVID-19 Vaccine Update survey prompted participants to record their vaccination status and, if applicable, answer questions regarding their vaccine. For this analysis, we included eligible AWHS participants who completed the COVID-19 Vaccine Update survey and tracked at least one menstrual cycle.

### COVID-19 Vaccine Update survey and cycle tracking

The COVID-19 Vaccine Update survey was first delivered to participants in September 2021. This survey was redistributed quarterly or until participants completed it. It asked if participants received a COVID-19 vaccine, and they could respond “Yes,” “No,” or “Prefer not to answer.” Participants who answered “Yes” received follow-up questions to ascertain the date of the first and second (if applicable) vaccines, the type of first and second (if applicable) vaccines (Pfizer-BioNTech, Moderna, Johnson & Johnson/Janssen, or other), and any symptoms in the 48 hours following a dose. Participants who completed the COVID-19 Vaccine Update survey with missing or incorrect dates [e.g., vaccination dates recorded before December 2021 (when vaccinations first became available) or after February 2022 (when this analysis was conducted) or second dose recorded before first dose] were excluded from this analysis. Dissemination of the COVID-19 Vaccine Update survey occurred prior to the authorization and roll out of additional doses and boosters in the U.S..^27^ Therefore, questions concerning the number of vaccines received was aligned with CDC classification of vaccination primary series.^28^

Characteristics of tracked menstrual cycles in the AWHS and methods for menstrual cycle identification have been described.^29^ Participants logged menses, with associated dates, via Cycle Tracking in the Apple Health app or any third-party cycle tracking app that the participant permitted to write to HealthKit. These merged data could include logging collected prospectively after enrollment and entries from up to two years prior to enrollment. Menstrual cycle length was derived from cycles identified after removing artifacts due to skipped tracking.^29^ The primary analysis included cycles tracked retrospectively as early as January 2018, and prospectively between enrollment and November 2021 when participants had not reported a hysterectomy and were not pregnant, lactating, menopausal, or using hormones.

For the current work, participants were categorized as vaccinated or unvaccinated as self-reported in the COVID-19 Vaccine Update survey. Of the participants who responded “Prefer not to answer” concerning their vaccination status (N = 216) and participants who indicated that they had received one dose of a two-dose series (N = 257), none tracked menstrual cycles. Individual cycles for vaccinated participants were labeled as: 1) “pre-vaccination”, if the entire cycle occurred prior to the first vaccination, 2) “first dose,” cycle in which the participant received the first vaccine dose of the Pfizer-BioNTech or Moderna series (i.e., mRNA vaccines), 3) “second dose,” the cycle in which the participant received the second dose of an mRNA series, 4) “J&J dose,” the cycle in which the participant received the single Johnson & Johnson/Janssen dose, or 5) “post-vaccination,” with the number of cycles since vaccination (since the second dose of an mRNA series or since the single J&J dose, accordingly) recorded. Post-vaccine cycles 1-4 were assessed for potentially persistent associations over time; we limited analysis to four cycles post-vaccination because booster vaccines likely became available as early as cycle five. We further categorized vaccination as having occurred during the follicular phase or luteal phase of a cycle to assess the impact of timing of the dose within a menstrual cycle. For a given cycle beginning with the first bleed day, we defined the luteal phase as the 14 days prior to the start of the subsequent cycle, and we defined the follicular phase as the remaining days beginning with the first bleed day and ending on the day prior to the luteal phase.^30, 31^ Participants with a single cycle in which they received both doses were excluded from the current analysis due to the fact that these cycles could not be short, by definition, and their length was not necessarily a result of treatment.

### Covariates

Extensive data were collected from survey questionnaires through the Apple Research app.^26^ Final regression models included covariates that were a priori defined as potential confounders based on previous literature and a directed acyclic graph [(DAG) Supplemental Figure S1].^32, 33^ These include age, race/ethnicity, body mass index (BMI), parity, and season. We calculated participants’ age as the year of a given menstrual cycle minus their birth year. Because previous studies have suggested a non-linear relationship between age and menstrual cycle length,^30, 34^ we categorized age as < 20, 20-24, 25-29, 30-34, 35-39, 40-44, 45-49, and > 50. For this analysis, we divided race/ethnicity into three categories based on self-report at enrollment, White, non-Hispanic, Hispanic (Latina or Spanish), or other [including Black, non-Hispanic (African American or African), Asian, American Indian or Alaska Native, Middle Eastern or North African, Native Hawaiian or Pacific Islander, more than one race/ethnicity, or participants who reported that none of the available categories fully described them], due to the small number of participants in some groups. BMI was calculated from self-reported height and weight at enrollment; participants are prompted to update these fields yearly. Participants were categorized as underweight (BMI < 18.5 kg/m^2^), normal (18.5 ≤ BMI < 25 kg/m^2^), overweight (25 ≤ BMI < 30 kg/m^2^), or obese (BMI ≥ 30 kg/m^2^). Parity was recorded as nulliparous or parous as reported in the reproductive history survey. We included an indicator variable for month and year of cycle to control for seasonality as well as shared pandemic experiences and stressors among all participants in the study (vaccinated and unvaccinated). Missingness was included as a category for all covariates.

### Statistical analysis

We examined distributional plots, contingency tables, and descriptive statistics for all variables of interest. We used conditional linear regression models with subject-level fixed effects to estimate the covariate-adjusted within-woman difference and 95% confidence intervals (95% CIs) in mean cycle length, measured in days, between pre-vaccination cycles and cycles in which a vaccine dose was administered and between pre-vaccination cycles and post-vaccination cycles. We included participant ID as a fixed effect in the conditional model to account for longitudinal clustering of repeated measures within each participant. The fixed effect controls for all potential confounders that vary between participants, and so in this approach, subject-specific covariates that do not vary with time are not included in the model. This strategy is similar to a case-control study that includes matching of cases and controls on certain covariates. In the present analysis, the “matching” is within a participant and compares vaccinated and post-vaccine cycles (case cycles) to their own pre-vaccine cycles (control cycles).

We defined long (>38 days) cycles using recommendations from the International Federation of Gynecology and Obstetrics and estimated the probability of experiencing a long menstrual cycle in cycles in which a vaccine was received and post-vaccine cycles compared with pre-vaccine cycles.^35^ To consider the chance of a long cycle in relation to vaccination, we fit a covariate-adjusted conditional logistic regression and present odds ratios (ORs) and 95% CIs.

We further compared associations between vaccination and menstrual cycle length by the timing of vaccine dose within a menstrual cycle (i.e., in the follicular phase or the luteal phase). Heterogeneity by menstrual cycle phase was considered by calculating the difference in effect estimates (e.g., first dose in the follicular phase versus first dose in the luteal phase) with the associated 95% CI; a 95% CI that did not contain zero was interpreted as a significant difference between the two coefficients.

Because conditional regression does not provide estimates for independent variables that do not vary within participant (e.g., race/ethnicity and parity), we used multivariable linear mixed-effects models with random participant-specific intercepts to estimate the covariate-adjusted difference and 95% CI in mean cycle length between vaccinated and unvaccinated participants. Additionally, we used generalized estimating equations (GEE) to fit a logistic regression model that estimated ORs and 95% CI for the association between COVID-19 vaccination and the probability of having a long cycle in vaccinated participants compared with unvaccinated participants.

We conducted additional sensitivity analyses to assess the robustness of our results. First, we restricted to participants who contributed at least three menstrual cycles; for vaccinated participants, we required at least two cycles before their first dose and at least one cycle in which a dose was administered or post-vaccination. Within participants who contributed three or more cycles, we calculated the average menstrual cycle length across all tracked cycles for unvaccinated participants and across pre-vaccination cycles for vaccinated participants and then restricted to those with an average cycle length of 24-38 days. Additionally, we retained participants who received two doses in a single cycle (N = 830), and we conducted a complete case analysis by removing cycles with missing values for covariates. Finally, considering some participants may have reported being unvaccinated in the COVID-19 Vaccine Update survey but later received a vaccine, we restricted to cycles recorded prior to COVID-19 Vaccine Update survey completion. Data management, processing, and statistical analyses were conducted in Python 3.6.^36^ All statistical tests were two-sided. We present Bonferroni-adjusted 95% confidence intervals for all results, with α = 0.05/7 (i.e., 99.99% confidence intervals) for the main analysis to account for multiple comparisons among the seven main associations of interest (i.e., first dose, second dose, J&J dose, and post-vaccination cycles 1-4, all compared with pre-vaccination cycles).

## Results

A total of 257,838 menstrual cycles from 15,001 participants who completed the COVID-19 Vaccine Update survey and tracked at least one cycle were initially identified. After applying the remaining exclusion criteria, 128,094 menstrual cycles from 9,652 participants were included in the current analysis (Supplemental Figure S2). Participants recorded 13 cycles on average [median = 10; inter-quartile range (IQR): 4-22 (Supplemental Figure S3)]. The average within-individual standard deviation in cycle length was 4.2 days. Eighty-eight percent of participants were vaccinated; 12% were not. Fifty-five percent of vaccinated participants received the Pfizer-BioNTech vaccine, 37% received Moderna, and 7% received Johnson & Johnson/Janssen. A total of 7,490 long cycles were identified, 6% of all cycles included. Vaccinated participants tended to be in the older age groups, to have a college or graduate degrees, to be married, and to be nulliparous (Table 1). The distribution of BMI and race/ethnicity were similar between vaccinated and unvaccinated participants.

**Table 1:** Demographic characteristics among 9,652 participants in the Apple Women’s Health Study at enrollment and number of cycles included among 128,094 total cycles

|  | Vaccinated<sup>a</sup> |  | Unvaccinated<sup>a</sup> |  | Overall<sup>a</sup> |  |
| --- | --- | --- | --- | --- | --- | --- |
|  | Participants | Cycles | Participants | Cycles | Participants | Cycles |
|  | (N<sub>p</sub> = 8,486)<sup>b</sup> | (N<sub>c</sub> = 113,890)<sup>b</sup> | (N<sub>p</sub> = 1,166)<sup>b</sup> | (N<sub>c</sub> = 14,204)<sup>b</sup> | (N<sub>p</sub> = 9,652)<sup>b</sup> | (N<sub>c</sub> = 128,094)<sup>b</sup> |
| Age |  |  |  |  |  |  |
| < 20 | 240 (2.8) | 11843 (10.4) | 58 (5.0) | 1724 (12.1) | 298 (3.1) | 13567 (10.6) |
| 20-24 | 1038 (12.2) | 16907 (14.8) | 168 (14.4) | 2294 (16.2) | 1206 (12.5) | 19201 (15.0) |
| 25-29 | 1475 (17.4) | 19345 (17.0) | 220 (18.9) | 2974 (20.9) | 1695 (17.6) | 22319 (17.4) |
| 30-34 | 1601 (18.9) | 22454 (19.7) | 259 (22.2) | 2904 (20.4) | 1860 (19.3) | 25358 (19.8) |
| 35-39 | 1606 (18.9) | 21619 (19.0) | 219 (18.8) | 2172 (15.3) | 1825 (18.9) | 23791 (18.6) |
| 40-44 | 1361 (16.0) | 14175 (12.4) | 144 (12.3) | 1137 (8.0) | 1505 (15.6) | 15312 (12.0) |
| 45-49 | 848 (10.0) | 3210 (2.8) | 75 (6.4) | 672 (4.7) | 923 (9.6) | 3882 (3.0) |
| >= 50 | 317 (3.7) | 4337 (3.8) | 23 (2.0) | 327 (2.3) | 340 (3.5) | 4664 (3.6) |
| Race/Ethnicity<sup>c</sup> |  |  |  |  |  |  |
| White, non-Hispanic | 6276 (74.0) | 84242 (74.0) | 848 (72.7) | 10334 (72.8) | 7124 (73.8) | 94576 (73.8) |
| Hispanic/Latina | 910 (10.7) | 11651 (10.2) | 137 (11.7) | 1559 (11.0) | 1047 (10.8) | 13210 (10.3) |
| Other | 1300 (15.3) | 17997 (15.8) | 181 (15.5) | 2311 (16.3) | 1481 (15.3) | 20308 (15.9) |
| Educational Attainment |  |  |  |  |  |  |
| No College Degree | 2854 (33.6) | 36086 (31.7) | 875 (75.0) | 10329 (72.7) | 3729 (38.6) | 46415 (36.2) |
| College Degree | 3081 (36.3) | 42715 (37.5) | 200 (17.2) | 2643 (18.6) | 3281 (34.0) | 45358 (35.4) |
| Graduate Degree | 2533 (29.8) | 34935 (30.7) | 84 (7.2) | 1204 (8.5) | 2617 (27.1) | 36139 (28.2) |
| Missing | 18 (0.2) | 154 (0.1) | 7 (0.6) | 28 (0.2) | 25 (0.3) | 182 (0.1) |
| Marital Status |  |  |  |  |  |  |
| Married | 3999 (47.1) | 54718 (48.0) | 433 (37.1) | 5435 (38.3) | 4432 (45.9) | 60153 (47.0) |
| Never married | 2602 (30.7) | 35478 (31.2) | 372 (31.9) | 4271 (30.1) | 2974 (30.8) | 39749 (31.0) |
| Widowed, divorced, or separated | 755 (8.9) | 10128 (8.9) | 180 (15.4) | 2392 (16.8) | 935 (9.7) | 12520 (9.8) |
| Part of an unmarried couple | 1098 (12.9) | 13120 (11.5) | 174 (14.9) | 2020 (14.2) | 1272 (13.2) | 15140 (11.8) |
| Missing | 32 (0.4) | 446 (0.4) | 7 (0.6) | 86 (0.6) | 39 (0.4) | 532 (0.4) |
| BMI<sup>d</sup> |  |  |  |  |  |  |
| Underweight | 139 (1.6) | 1880 (1.7) | 43 (3.7) | 587 (4.1) | 182 (1.9) | 2467 (1.9) |
| Normal weight | 2899 (34.2) | 39840 (35.0) | 355 (30.4) | 4474 (31.5) | 3254 (33.7) | 44314 (34.6) |
| Overweight | 2192 (25.8) | 28828 (25.3) | 300 (25.7) | 3659 (25.8) | 2492 (25.8) | 32487 (25.4) |
| Obese | 3108 (36.6) | 39800 (34.9) | 450 (38.6) | 5263 (37.1) | 3558 (36.9) | 45063 (35.2) |
| Missing | 148 (1.7) | 3542 (3.1) | 18 (1.5) | 221 (1.6) | 166 (1.7) | 3763 (2.9) |
| Parity |  |  |  |  |  |  |
| Parous | 3335 (39.3) | 45251 (39.7) | 643 (55.1) | 7951 (56.0) | 3978 (41.2) | 53202 (41.5) |
| Nulliparous | 5101 (60.1) | 67986 (59.7) | 512 (43.9) | 6200 (43.6) | 5613 (58.2) | 74186 (57.9) |
| Missing | 50 (0.6) | 653 (0.6) | 11 (0.9) | 53 (0.4) | 61 (0.6) | 706 (0.6) |

<sup>a</sup> Values are presented as N (%) of total participants included in this analysis
<sup>b</sup> N<sub>p</sub> = number of participants. N<sub>c</sub> = number of cycles
<sup>c</sup> Including Black, non-Hispanic (African American or African), Asian, American Indian or Alaska Native, Middle Eastern or North African, Native Hawaiian or Pacific Islander, more than one race/ethnicity, or participants who reported that none of the available categories fully described them
<sup>d</sup> BMI, body mass index. Underweight: BMI < 18.5 kg/m<sup>2</sup>; normal: 18.5 ≤ BMI < 25 kg/m<sup>2</sup>, overweight: 25 ≤ BMI < 30 kg/m<sup>2</sup>, Obese: BMI ≥ 30 kg/m<sup>2</sup>

Using covariate-adjusted conditional linear regression, we found menstrual cycles when participants received the vaccine to be, on average, longer than their pre-vaccination cycles (Table 2). Among vaccinated participants, COVID-19 vaccination was associated with a small increase in length for cycles when participants received the first dose (0.50 days, 95% CI: 0.22, 0.78) and cycles when participants received the second dose (0.39 days, 95% CI: 0.11, 0.67) of mRNA vaccines compared with pre-vaccination cycles. Cycles in which the single dose of J&J was administered were, on average, 1.26 days longer (95% CI: 0.45, 2.07) than pre-vaccination cycles. Post-vaccination cycles returned to average pre-vaccination length, with 0.14 days (95% CI: −0.13, 0.40) in the first cycle following vaccination, 0.13 days (95% CI: −0.14, 0.40) in the second cycle, −0.17 days (95% CI: −0.43, 0.10) in the third cycle, and −0.25 days (95% CI: −0.52, 0.01) in the fourth.

**Table 2.** Adjusted within-participant change in mean menstrual cycle length and 95% confidence intervals (95% CIs) and adjusted odds ratios (ORs) and 95% CIs of experiencing a long menstrual cycle (> 38 days) comparing cycles in which a vaccine was administered and post-vaccination cycles with pre-vaccination cycles

|  | Difference in mean cycle length (95% CI)<sup>a, b</sup> | Odds Ratio of having a long cycle (95% CI)<sup>a, c</sup> |
| --- | --- | --- |
| Pre-vaccination cycles | Reference | Reference |
| First dose<sup>d</sup> | 0.50 (0.22, 0.78) | 1.13 (0.86, 1.48) |
| Second dose<sup>d</sup> | 0.39 (0.11, 0.67) | 1.08 (0.82, 1.42) |
| J&J dose<sup>e</sup> | 1.26 (0.45, 2.07) | 2.17 (1.16, 4.04) |
| Post-vaccination cycles |  |  |
| First cycle | 0.14 (-0.13, 0.40) | 0.94 (0.73, 1.21) |
| Second cycle | 0.13 (-0.14, 0.40) | 0.94 (0.73, 1.21) |
| Third cycle | -0.17 (-0.43, 0.10) | 0.76 (0.59, 0.98) |
| Fourth cycle | -0.25 (-0.52, 0.01) | 0.71 (0.55, 0.93) |

<sup>a</sup> Conditional models control for all participant-level characteristics; models additionally adjusted for age, BMI, and seasonality
<sup>b</sup> Differences are from conditional linear regression
<sup>c</sup> Odds ratios are from conditional logistic regression
<sup>d</sup> Doses 1 and 2 were Pfizer-BioNTech or Moderna
<sup>e</sup> J&J = Johnson & Johnson/Janssen

The conditional logistic regression model of the probability of a long cycle suggested that, compared with pre-vaccination cycles, participants were more likely to experience a long cycle during the cycle in which they received the J&J vaccine [OR = 2.17, 95% CI: 1.16, 4.04 (Table 2)]. There was no evidence of increased probability of a long cycle for cycles in which participants received the first dose (OR = 1.13, 95% CI: 0.86, 1.48) or the second dose (OR = 1.08, 95% CI: 0.82, 1.42) of mRNA vaccines. There was, likewise, no evidence of increased odds of long cycles in the first cycle (OR = 0.94, 95% CI: 0.73, 1.21) or the second cycle (OR = 0.94, 95% CI: 0.73, 1.21) post-vaccination, regardless of the vaccine type. In the third and fourth post-vaccination cycles, the odds of a long cycle decreased, with OR = 0.76 (95% CI: 0.59, 0.98) in the third cycle and OR = 0.71 (95% CI: 0.55, 0.93) in the fourth cycle following the single J&J dose or the second dose of an mRNA vaccine.

The association between vaccine dose and mean cycle length depended on the phase in which the dose was received (Table 3). Compared with pre-vaccination cycles, follicular phase vaccination with an mRNA vaccine was associated with an increase in mean cycle length in first dose cycles (0.97 days, 95% CI: 0.53, 1.42) or second dose cycles (1.43 days, 95% CI: 1.06, 1.80). For the J&J dose, follicular phase vaccination was associated with a 2.27 (95% CI: 1.04, 3.50) day increase in cycle length. There was no evidence of increased mean cycle length in cycles in which the first vaccine dose (0.21 days, 95% CI: −0.14, 0.57) or the J&J vaccine dose (0.39 days, 95% CI: −0.75, 1.53) were administered in the luteal phase. However, luteal phase vaccination was associated with a decrease in average length for cycles in which the second dose was administered (−0.97 days, 95% CI: −1.39, −0.55), compared with pre-vaccination cycles. Regression coefficients for post-vaccination cycles did not meaningfully change with the inclusion of cycle phase. In the covariate-adjusted mixed-effects model, we found no evidence of a difference between mean menstrual cycle length in vaccinated and unvaccinated participants (0.24 days, 95% CI: −0.34, 0.82). Likewise, in the GEE, the probability of a long cycle was similar in unvaccinated and vaccinated participants (OR = 1.20, 95% CI: 1.00, 1.44). Regression coefficients for vaccine doses were similar to but larger in magnitude than those from conditional regression models (Tables S1 and S2).

**Table 3.** Adjusted within-participant change in mean menstrual cycle length and 95% confidence intervals (95% CIs) comparing cycles in which a vaccine was administered and post-vaccination cycles with pre-vaccination cycles by menstrual cycle phase of vaccine dose

|  | Difference in mean cycle length (95% CI)<sup>a, b</sup> | Difference from coefficient for dose in luteal phase (95% CI)<sup>a, c</sup> |
| --- | --- | --- |
| Pre-vaccination cycles | Reference | --- |
| Dose in follicular phase |  |  |
| First dose<sup>d</sup> | 0.97 (0.53, 1.42) | 0.76 (0.16, 1.35) |
| Second dose<sup>d</sup> | 1.43 (1.06, 1.80) | 2.40 (1.81, 2.99) |
| J&J dose<sup>e</sup> | 2.27 (1.04, 3.50) | 1.89 (0.20, 3.27) |
| Dose in luteal phase |  |  |
| First dose<sup>d</sup> | 0.21 (-0.14, 0.57) | Reference |
| Second dose<sup>d</sup> | -0.97 (-1.39, -0.55) | Reference |
| J&J dose<sup>e</sup> | 0.39 (-0.75, 1.53) | Reference |
| Post-vaccination cycles |  |  |
| First cycle | 0.16 (-0.11, 0.44) | --- |
| Second cycle | 0.15 (-0.12, 0.43) | --- |
| Third cycle | -0.14 (-0.42, 0.13) | --- |
| Fourth cycle | -0.24 (-0.51, 0.04) | --- |

<sup>a</sup> Conditional model controls for all participant-level characteristics; model additionally adjusted for age, BMI, and seasonality
<sup>b</sup> Differences are from conditional linear regression
<sup>c</sup> Difference between regression coefficients for a given dose in the follicular vs luteal phase
<sup>d</sup> Doses 1 and 2 were Pfizer-BioNTech or Moderna
<sup>e</sup> J&J = Johnson & Johnson/Janssen

Fifty-eight percent of participants were included in the sensitivity analysis restricted to those with three or more tracked cycles. While 82% of participants had three or more tracked cycles, only 53% of vaccinated participants had two or more tracked cycles before vaccination and at least one tracked cycle in which a vaccine was administered or after vaccination. Three percent of cycles had missing values and were dropped in the complete case analysis. Ninety-seven percent of cycles were tracked before completion of the COVID-19 Vaccine Update survey, and 83% of participants had an average cycle length of 24-38 days. Results from sensitivity analyses were consistent with those from the main analyses (Table S1-S4). When participants who received two doses in a single menstrual cycle were included, these cycles were, on average, 4.96 days (95% CI: 4.42, 5.50) longer than pre-vaccination cycles (Table S5).

## Discussion

In this large prospective cohort of participants in the U.S., we examined differences in menstrual cycle length in relation to the type of COVID-19 vaccine administered and the timing of administration. Our results suggest a small, non-persistent increase in average length of menstrual cycles in which a vaccine was administered. Moreover, the J&J vaccine was associated with a significant increase in the probability of a clinically long (> 38 days) cycle; however, this increase did not persist over time. We found little evidence of increased length for cycles following vaccination. We saw that doses received in the follicular phase explained the increase in mean cycle length observed in the primary results. We also observed that when a second dose was received in the luteal phase, that cycle was, on average, shorter. Together, these results indicate that the association between vaccination and cycle length differs by the phase in which a dose was administered.

Previous studies have shown increased cycle length,^20^ as well as menstrual cycle irregularities in relation to COVID-19 vaccination.^21–24^ The current work is consistent with results from Edelman et al. that showed a less than one day increase in length during cycles when vaccination occurred.^20^ We extended this work to evaluate vaccine timing and menstrual cycle length and found follicular dosing of all vaccines associated with longer cycles, while a second dose of an mRNA vaccine in the luteal phase was associated with slightly shorter cycles.

Potential mechanisms underlying the change in menstrual cycle length may involve inflammation from the immune response to vaccination. This immune response may impact a) signaling between the hypothalamus, pituitary, and ovaries (HPO), resulting in i) prolongation of follicular recruitment and, as a result, elongation of menstrual cycle length,^37^ or ii) suppression of the growth of the endometrial lining,^38^ and b) endometrial stability in the luteal phase, causing a reduction in cycle length.^39, 40^

The onset of menstruation is characterized by endometrial breakdown and apoptosis. Immune cells derived from menstrual blood demonstrate production of inflammatory markers (e.g., interferon (IFN)-gamma, granzyme B, and perforin),^41^ and genes related to inflammatory processes (e.g., lipopolysaccharide binding protein, SERPINB3, SERPINB4, IL17C, and SPINK1) are up-regulated.^42^ mRNA vaccines may potentially trigger an innate immune response by activating pro-inflammatory cytokines, type I IFNs, and pro-inflammatory genes.^43^ In this way, the Pfizer-BioNTech and Moderna vaccines may disturb the menstrual cycle by promoting local and systemic inflammation.

Individual cycle length varies naturally,^44, 45^ and the International Federation of Gynecology and Obstetrics classifies a difference of less than 8 days between an individual’s shortest and longest cycles as normal.^35^ The magnitude of the change we see in our data following vaccination is well within this natural variability. In contrast, infection by the COVID-19 virus may cause potentially long-term disruption of menstrual function.^46–51^ Research has found no evidence that COVID-19 vaccination adversely impacts fertility outcomes.^12–18^ COVID-19 vaccination is widely regarded as safe for people who menstruate,^52, 53^ and this work supports that conclusion.

This study has several strengths. First, a large sample size (120,815 menstrual cycles from 9,295 participants) allowed for sufficient statistical power to detect minor differences measured in fractions of days. Further, the AWHS includes a diverse population across a wide range of demographic characteristics (e.g., age, race/ethnicity, education, BMI category). Moreover, the AWHS is not limited to users of a specific cycle tracking app which may target certain demographic groups; instead, cycle tracking can occur directly through the Health app or through a variety of third-party apps. Additionally, menstrual cycle characteristics were reported prospectively. We used multivariable models to adjust for potential confounding in all analyses. In our main analyses, we used conditional, fixed effect models which control for individual-level features. This removed any avenue for confounding by sociodemographic factors or differential tracking behavior across participants. Finally, we controlled for seasonality by including an indicator variable for month and year of cycle; this further accounted for pandemic era experiences and shared social stressors in both vaccinated and unvaccinated participants.

Our findings should be interpreted in light of several limitations. First, because our analysis considered mean cycle length in the population, it cannot distinguish subpopulations, such as participants with preexisting conditions, who may have experienced more extreme changes in cycle length. Second, we relied on self-report for menstrual characteristics, COVID-19 vaccination details, and reported covariates. Consequently, the accuracy of the estimated menstrual cycle length may be affected by user tracking behavior, on which we have limited information. However, use of conditional models for our primary analyses controlled for differences in tracking behavior between participants, and the precise indicator variable for month and year controlled for potential changes in tracking behavior over time, leaving the only path for temporal confounding through differential changes in tracking behavior over time by treatment (i.e., in vaccinated and unvaccinated participants). We also had limited data on menses and were thus unable to address potential associations between the COVID-19 vaccine and bleed length or flow characteristics. We lacked measurements for participants’ hormone levels to determine the follicular and luteal phases of a given cycle; instead, we calculated this using a standard approach which could affect the phase-specific estimates and the comparisons between them.^30, 31^ Because our study population only included iPhone users in the U.S., our results may not be generalizable to all U.S. individuals who menstruate or to other populations. Additionally, given available data, we did not include participants with two vaccine doses in a single cycle in our main analyses. In a sensitivity analysis, these cycles appeared close to five days longer than pre-vaccination cycles, on average. However, cycles in which two vaccines were administered could not be shorter than the number of days between doses (usually 21 or 28 days), truncating the distribution of menstrual cycle length and making these cycles longer, by definition, and not necessarily a result of vaccination. Because receiving two doses in a single cycle was dependent on menstrual cycle length, we did not present these findings as main results, and further research is required to investigate this association. Finally, we cannot exclude the possibility of residual confounding from an unidentified time-varying factor, such as COVID-19 infection, which we did not query. Nonetheless, we adjusted for multiple factors that varied within participant (and between participants in mixed-effects models) to account for confounding from multiple directions. We do not expect, therefore, that residual confounding can fully explain our results.

## Conclusions

In the present study, we estimated the association between COVID-19 vaccination and menstrual cycle length in the cycles in which vaccination occurred and in succeeding cycles. Vaccination was associated with a very small increase in cycle length that was well within the natural variability in the study population. This change appeared driven by doses received in the follicular phase of the cycle. The magnitude of the increase diminished in each cycle following vaccination, and no association with cycle length persisted over time. This change appears minor and temporary and should not discourage individuals from becoming vaccinated.

## Data Availability

Aggregated data that support the findings of this study may be available upon request from the corresponding author SM. Any request for data will be evaluated and responded to in a manner consistent with policies intended to protect participant confidentiality and language in the Study protocol and informed consent form.

## Acknowledgements

The AWHS team would like to thank the participants for signing up for the study and contributing to the advancement of women’s health research. We would like to acknowledge Alexis de Figueiredo Veiga, MS, Nicola Gallagher, MS, and Ariel L Scalise, MPH for their work in supporting the study as part of the Harvard Study staff.

## Supplemental Materials

**Figure S1.**
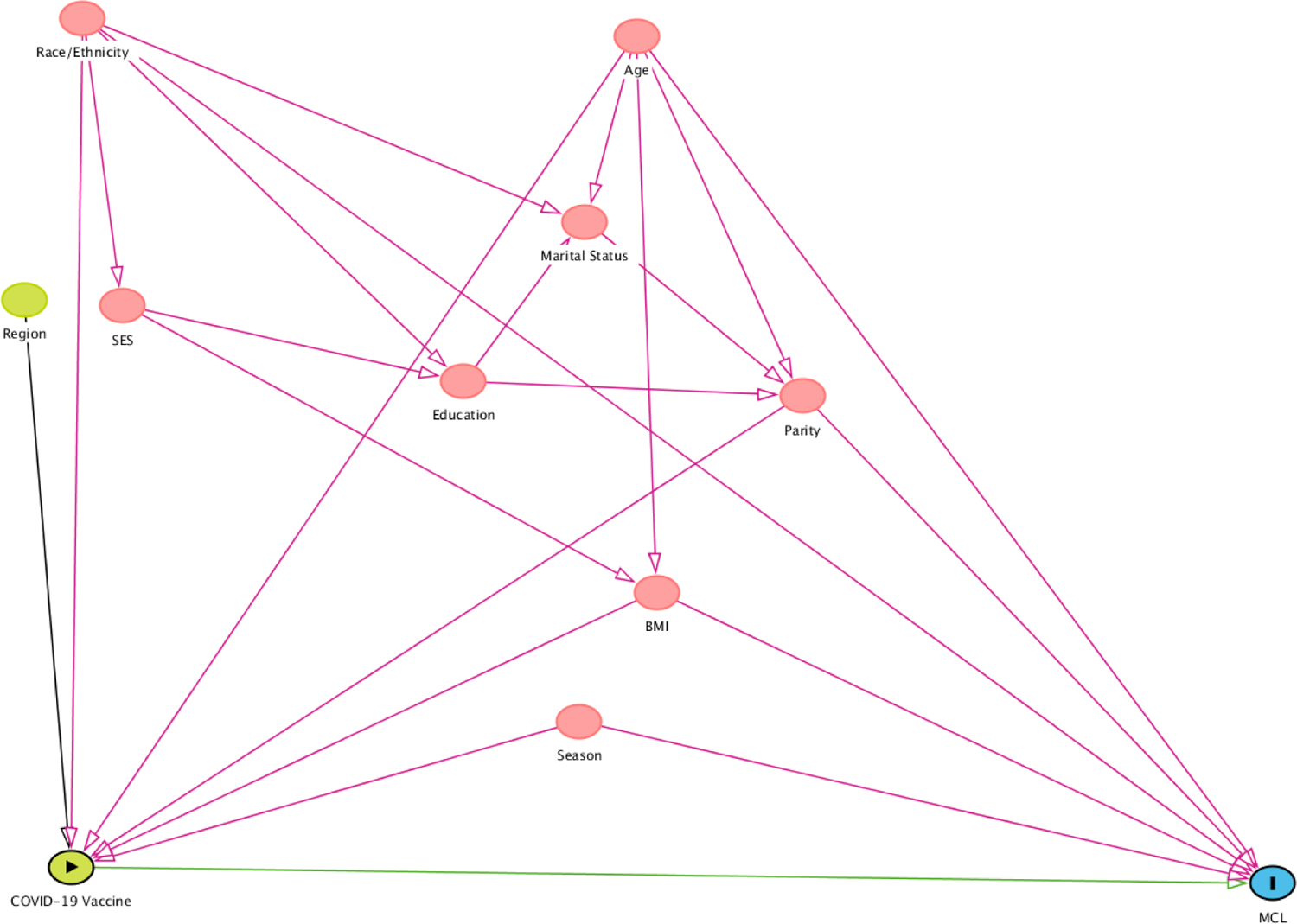
Directed Acyclic Graph (DAG) used to identify confounding variables as covariates for inclusion in regression models; Note: Figure generated using DAGitty v3.0.5. Acronyms: SES = socioeconomic status; BMI = body mass index; MCL = menstrual cycle length. The green variable with a triangle, COVID-19 vaccine, is the treatment, and the blue variable, MCL, is the outcome of interest. The green arrow is the association of interest. Variables in red (race/ethnicity, SES, education, age, marital status, BMI, parity, and season) are identified as confounders, and red arrows are confounding paths. The variable in green, region, affects only the exposure, and the black arrow is a non-confounding path. This DAG identifies five variables—age, race/ethnicity, parity, BMI, and season—as the minimally sufficient adjustment set for estimating the total effect of the COVID-19 vaccine on MCL.

**Figure S2.**
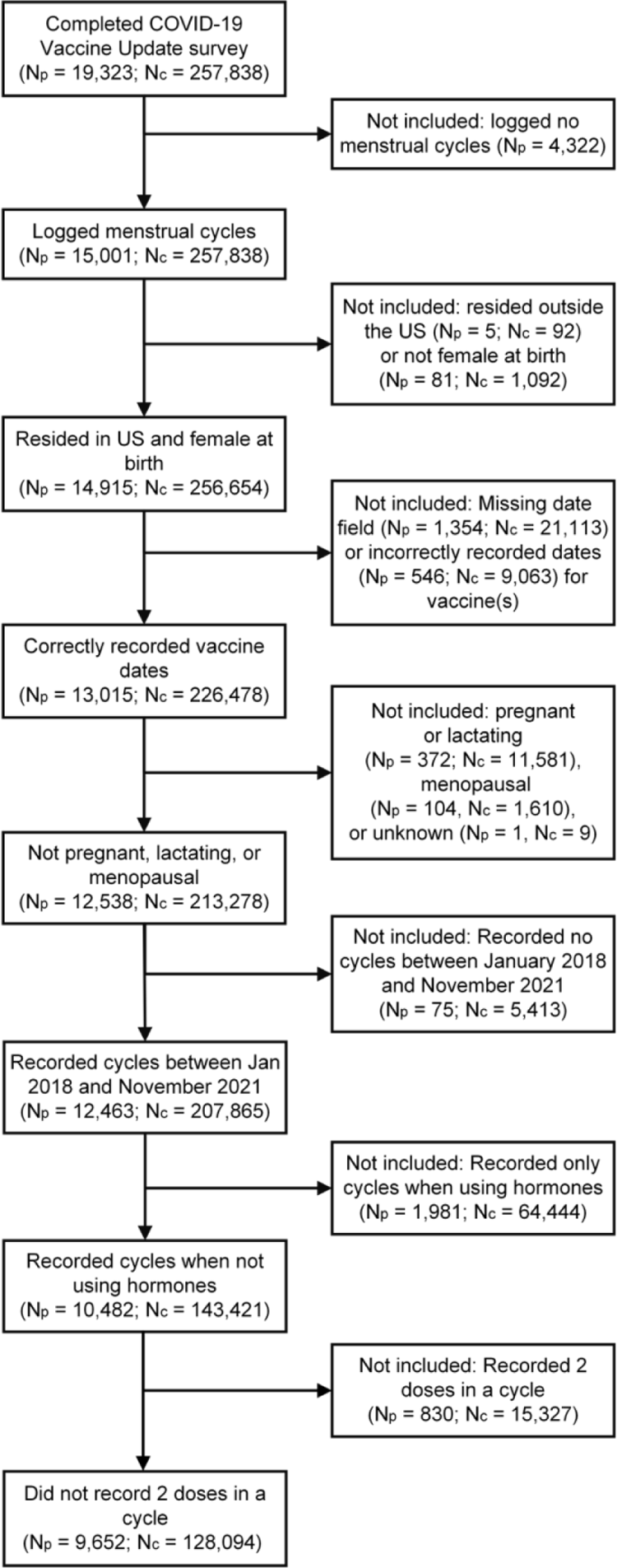
Flow chart of study identification, inclusion, and exclusion criteria for current analysis. N_p_ = number of participants. N_c_ = number of cycles.

**Figure S3.**
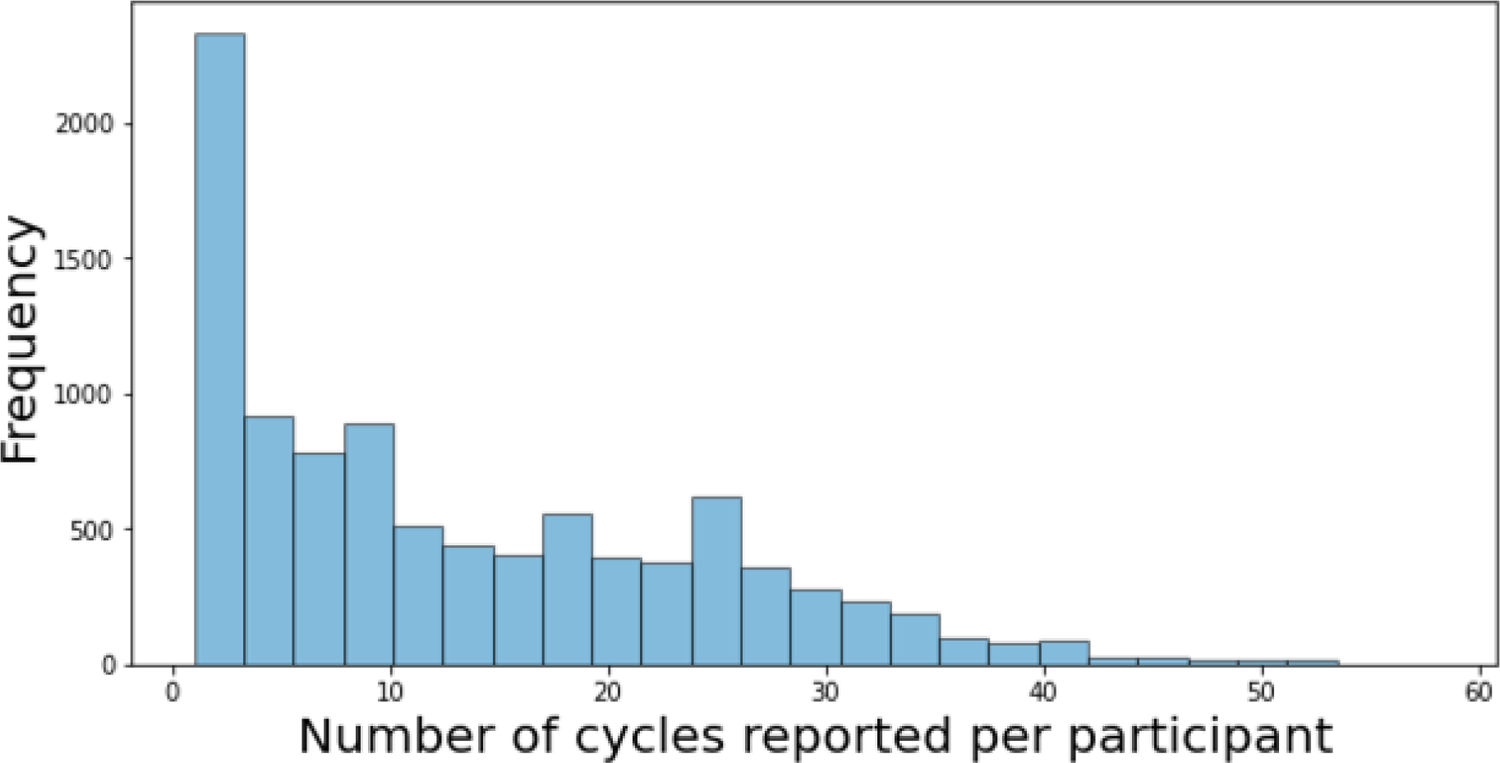
Histogram of number of cycles reported by participants in the Apple Women’s Health Study.

**Table S1.** Adjusted change in mean menstrual cycle length and 95% confidence intervals (95% CIs) and adjusted odds ratios (ORs) and 95% CIs of experiencing a long menstrual cycle (> 38 days) comparing vaccinated and unvaccinated participants and cycles in which a vaccine was administered and post-vaccination cycles with pre-vaccination cycles

|  | Difference in mean<br>cycle length (95% CI)<sup>a, b</sup> | Odds Ratio of having a<br>long cycle (95% CI)<sup>a, c</sup> |
| --- | --- | --- |
| Cycles in unvaccinated participants | 0.24 (-0.34, 0.82) | 1.20 (1.00, 1.44) |
| Pre-vaccination cycles | Reference | Reference |
| First dose<sup>d</sup> | 0.61 (0.33, 0.90) | 1.30 (1.11, 1.51) |
| Second dose<sup>d</sup> | 0.54 (0.26, 0.83) | 1.36 (1.16, 1.58) |
| J&J dose<sup>e</sup> | 1.39 (0.57, 2.22) | 2.17 (1.47, 3.19) |
| Post-vaccination cycles |  |  |
| First cycle | 0.43 (0.16, 0.70) | 1.48 (1.28, 1.72) |
| Second cycle | 0.35 (0.08, 0.62) | 1.40 (1.20, 1.63) |
| Third cycle | 0.00 (-0.26, 0.27) | 1.14 (0.98, 1.33) |
| Fourth cycle | -0.17 (-0.44, 0.10) | 0.95 (0.81, 1.11) |

<sup>a</sup> Models adjusted for race/ethnicity, parity, age, BMI, and seasonality
<sup>b</sup> Differences are from linear mixed-effect regression with participant-specific random intercepts
<sup>c</sup> Odds ratios are from logistic generalized estimating equations
<sup>d</sup> Doses 1 and 2 were Pfizer-BioNTech or Moderna
<sup>e</sup> J&J = Johnson & Johnson/Janssen

**Table S2.** Adjusted change in mean menstrual cycle length and 95% confidence intervals (95% CIs) comparing vaccinated and unvaccinated participants and cycles in which a vaccine was administered and post-vaccination cycles with pre-vaccination cycles

|  | Difference in mean cycle length (95% CI)<sup>a, b</sup> | Difference from coefficient for dose in luteal phase (95% CI)<sup>a, b, c</sup> |
| --- | --- | --- |
| Cycles in unvaccinated participants | 0.25 (-0.35, 0.85) | --- |
| Pre-vaccination cycles | Reference | --- |
| Dose in follicular phase |  |  |
| First dose<sup>d</sup> | 1.09 (0.64, 1.54) | 0.77 (0.16, 1.37) |
| Second dose<sup>d</sup> | 1.66 (1.28, 2.03) | 2.57 (1.97, 3.18) |
| J&J dose<sup>e</sup> | 2.42 (1.17, 3.66) | 1.92 (0.21, 3.64) |
| Dose in luteal phase |  |  |
| First dose<sup>d</sup> | 0.32 (-0.03, 0.68) | Reference |
| Second dose<sup>d</sup> | -0.92 (-1.34, -0.49) | Reference |
| J&J dose<sup>e</sup> | 0.50 (-0.66, 1.65) | Reference |
| Post-vaccination cycles |  |  |
| First cycle | 0.46 (0.18, 0.74) | --- |
| Second cycle | 0.37 (0.09, 0.65) | --- |
| Third cycle | 0.03 (-0.25, 0.30) | --- |
| Fourth cycle | -0.15 (-0.43, 0.13) | --- |

<sup>a</sup> Mixed-effect models adjusted for race/ethnicity, education, marital status, parity, age, BMI, and seasonality.
<sup>b</sup> Differences are from linear mixed-effect regression with participant-specific random intercepts
<sup>c</sup> Difference between regression coefficients for a given dose in the follicular vs luteal phase
<sup>d</sup> Doses 1 and 2 were Pfizer-BioNTech or Moderna
<sup>e</sup> J&J = Johnson & Johnson/Janssen

**Table S3.** Adjusted within-participant change in mean menstrual cycle length and 95% confidence intervals (95% CIs) comparing cycles in which a vaccine was administered and post-vaccination cycles with pre-vaccination cycles after restricting to participants who tracked at least 3 cycles, restricting to participants with average cycles between 24-38 days, restricting to cycles completed before COVID-19 Vaccine Update survey completion, and restricting to cycles with complete cases

|  | Participants with 3 or more cycles | Participants with normal average cycles | Cycles before COVID-19 survey completion | Cycles with complete cases |
| --- | --- | --- | --- | --- |
|  | Difference in mean cycle length (95% CI)<sup>a, b</sup> | Difference in mean cycle length (95% CI)<sup>a, b</sup> | Difference in mean cycle length (95% CI)<sup>a, b</sup> | Difference in mean cycle length (95% CI)<sup>a, b</sup> |
| Pre-vaccination cycles | Reference | Reference | Reference | Reference |
| First dose<sup>c</sup> | 0.52 (0.25, 0.79) | 0.48 (0.23, 0.73) | 0.51 (0.23, 0.79) | 0.49 (0.21, 0.78) |
| Second dose<sup>c</sup> | 0.43 (0.15, 0.71) | 0.41 (0.15, 0.67) | 0.39 (0.10, 0.67) | 0.38 (0.09, 0.67) |
| J&J dose<sup>d</sup> | 1.29 (0.51, 2.08) | 1.22 (0.48, 1.96) | 1.33 (0.52, 2.14) | 1.34 (0.52, 2.17) |
| Post-vaccination cycles |  |  |  |  |
| First cycle | 0.14 (-0.12, 0.41) | 0.12 (-0.13, 0.37) | 0.14 (-0.12, 0.41) | 0.15 (-0.12, 0.43) |
| Second cycle | 0.30 (0.02, 0.57) | 0.32 (0.06, 0.58) | 0.13 (-0.13, 0.40) | 0.15 (-0.12, 0.42) |
| Third cycle | 0.00 (-0.28, 0.28) | 0.01 (-0.25, 0.27) | -0.13 (-0.39, 0.14) | -0.13 (-0.40, 0.15) |
| Fourth cycle | -0.16 (-0.44, 0.13) | -0.06 (-0.32, 0.21) | -0.23 (-0.49, 0.04) | -0.25 (-0.52, 0.02) |

<sup>a</sup> Differences are from conditional linear regression
<sup>b</sup> Conditional model controls for all participant-level characteristics; model additionally adjusted for age, BMI, and seasonality
<sup>c</sup> Doses 1 and 2 were Pfizer-BioNTech or Moderna
<sup>d</sup> J&J = Johnson & Johnson/Janssen

**Table S4.**
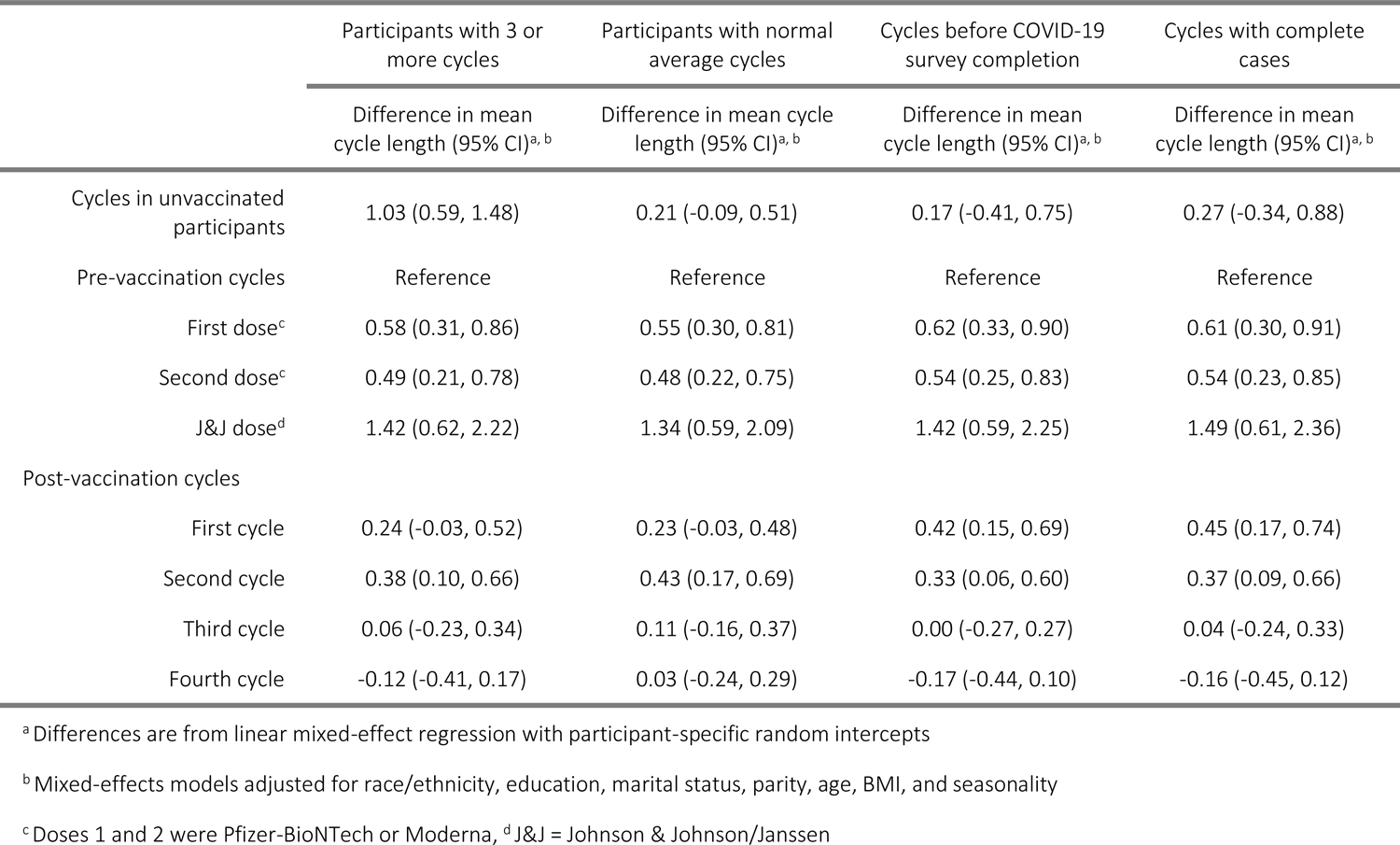
Adjusted change in mean menstrual cycle length and 95% confidence intervals (95% CIs) comparing vaccinated and unvaccinated participants and cycles in which a vaccine was administered and post-vaccination cycles with pre-vaccination cycles after restricting to participants who tracked at least 3 cycles, restricting to participants with average cycles between 24-38 days, restricting to cycles completed before COVID-19 Vaccine Update survey completion, and restricting to cycles with complete cases

|  | Participants with 3 or more cycles | Participants with normal average cycles | Cycles before COVID-19 survey completion | Cycles with complete cases |
| --- | --- | --- | --- | --- |
|  | Difference in mean cycle length (95% CI)<sup>a, b</sup> | Difference in mean cycle length (95% CI)<sup>a, b</sup> | Difference in mean cycle length (95% CI)<sup>a, b</sup> | Difference in mean cycle length (95% CI)<sup>a, b</sup> |
| Cycles in unvaccinated participants | 1.03 (0.59, 1.48) | 0.21 (-0.09, 0.51) | 0.17 (-0.41, 0.75) | 0.27 (-0.34, 0.88) |
| Pre-vaccination cycles | Reference | Reference | Reference | Reference |
| First dose<sup>c</sup> | 0.58 (0.31, 0.86) | 0.55 (0.30, 0.81) | 0.62 (0.33, 0.90) | 0.61 (0.30, 0.91) |
| Second dose<sup>c</sup> | 0.49 (0.21, 0.78) | 0.48 (0.22, 0.75) | 0.54 (0.25, 0.83) | 0.54 (0.23, 0.85) |
| J&J dose<sup>d</sup> | 1.42 (0.62, 2.22) | 1.34 (0.59, 2.09) | 1.42 (0.59, 2.25) | 1.49 (0.61, 2.36) |
| Post-vaccination cycles |  |  |  |  |
| First cycle | 0.24 (-0.03, 0.52) | 0.23 (-0.03, 0.48) | 0.42 (0.15, 0.69) | 0.45 (0.17, 0.74) |
| Second cycle | 0.38 (0.10, 0.66) | 0.43 (0.17, 0.69) | 0.33 (0.06, 0.60) | 0.37 (0.09, 0.66) |
| Third cycle | 0.06 (-0.23, 0.34) | 0.11 (-0.16, 0.37) | 0.00 (-0.27, 0.27) | 0.04 (-0.24, 0.33) |
| Fourth cycle | -0.12 (-0.41, 0.17) | 0.03 (-0.24, 0.29) | -0.17 (-0.44, 0.10) | -0.16 (-0.45, 0.12) |

<sup>a</sup> Differences are from linear mixed-effect regression with participant-specific random intercepts
<sup>b</sup> Mixed-effects models adjusted for race/ethnicity, education, marital status, parity, age, BMI, and seasonality
<sup>c</sup> Doses 1 and 2 were Pfizer-BioNTech or Moderna, <sup>d</sup> J&J = Johnson & Johnson/Janssen

**Table S5.** Adjusted within-participant change in mean menstrual cycle length and 95% confidence intervals (95% CIs) comparing cycles in which a vaccine was administered and post-vaccination cycles with pre-vaccination cycles, retaining participants who received two doses in a single cycle.

|  | Difference in mean cycle length (95% CI)<sup>a</sup> |
| --- | --- |
| Pre-vaccination cycles | Reference |
| First dose<sup>b</sup> | 0.51 (0.23, 0.79) |
| Second dose<sup>b</sup> | 0.42 (0.13, 0.70) |
| J&J dose<sup>c</sup> | 1.28 (0.46, 2.10) |
| First and second dose<sup>b</sup> | 4.96 (4.42, 5.50) |
| Post-vaccination cycles |  |
| First cycle | 0.18 (-0.07, 0.43) |
| Second cycle | 0.15 (-0.10, 0.41) |
| Third cycle | -0.11 (-0.36, 0.14) |
| Fourth cycle | -0.19 (-0.44, 0.06) |

<sup>a</sup> Conditional models control for all participant-level characteristics; models additionally adjusted for age, BMI, and seasonality
<sup>b</sup> Doses 1 and 2 were Pfizer-BioNTech or Moderna
<sup>c</sup> J&J = Johnson & Johnson/Janssen

STROBE Statement—Checklist of items that should be included in reports of *cohort studies*

|  | Item No | Recommendation | Page No |
| --- | --- | --- | --- |
| Title and abstract | 1 | (a) Indicate the study’s design with a commonly used term in the title or the abstract | 2 |
|  |  | (b) Provide in the abstract an informative and balanced summary of what was done and what was found | 2 |
| Introduction |  |  |  |
| Background/rationale | 2 | Explain the scientific background and rationale for the investigation being reported | 3 |
| Objectives | 3 | State specific objectives, including any prespecified hypotheses | 4 |
| Methods |  |  |  |
| Study design | 4 | Present key elements of study design early in the paper | 4-5 |
| Setting | 5 | Describe the setting, locations, and relevant dates, including periods of recruitment, exposure, follow-up, and data collection | 4-5 |
| Participants | 6 | (a) Give the eligibility criteria, and the sources and methods of selection of participants. Describe methods of follow-up | 4-5 |
|  |  | (b) For matched studies, give matching criteria and number of exposed and unexposed | NA |
| Variables | 7 | Clearly define all outcomes, exposures, predictors, potential confounders, and effect modifiers. Give diagnostic criteria, if applicable | 5-6 |
| Data sources/measurement | 8* | For each variable of interest, give sources of data and details of methods of assessment (measurement). Describe comparability of assessment methods if there is more than one group | 5-6 |
| Bias | 9 | Describe any efforts to address potential sources of bias | 6-7 |
| Study size | 10 | Explain how the study size was arrived at | 5-7, Fig S1 |
| Quantitative variables | 11 | Explain how quantitative variables were handled in the analyses. If applicable, describe which groupings were chosen and why | 5-7 |
| Statistical methods | 12 | (a) Describe all statistical methods, including those used to control for confounding | 7-8 |
|  |  | (b) Describe any methods used to examine subgroups and interactions | NA |
|  |  | (c) Explain how missing data were addressed | 7 |
|  |  | (d) If applicable, explain how loss to follow-up was addressed | NA |
|  |  | (e) Describe any sensitivity analyses | 8-9 |
| Results |  |  |  |
| Participants | 13* | (a) Report numbers of individuals at each stage of study—eg numbers potentially eligible, examined for eligibility, confirmed eligible, included in the study, completing follow-up, and analysed | 9, Fig S1 |
|  |  | (b) Give reasons for non-participation at each stage | 9, Fig S1 |
|  |  | (c) Consider use of a flow diagram | Fig S1 |
| Descriptive data | 14* | (a) Give characteristics of study participants (eg demographic, clinical, social) and information on exposures and potential confounders | 9, Table 1 |
|  |  | (b) Indicate number of participants with missing data for each variable of interest | 9, Table 1 |
|  |  | (c) Summarise follow-up time (eg, average and total amount) | 9, Fig S3 |
| Outcome data | 15* | Report numbers of outcome events or summary measures over time | 9 |
| Main results | 16 | (a) Give unadjusted estimates and, if applicable, confounder-adjusted estimates and their precision (eg, 95% confidence interval). Make clear which confounders were adjusted for and why they were included<br>(b) Report category boundaries when continuous variables were categorized<br>(c) If relevant, consider translating estimates of relative risk into absolute risk for a meaningful time period | 9-11,<br>Tables 2-3<br>6<br>NA |
| Other analyses | 17 | Report other analyses done—eg analyses of subgroups and interactions, and sensitivity analyses | 11,<br>Tables S1-S4 |
| **Discussion** |  |  |  |
| Key results | 18 | Summarise key results with reference to study objectives | 11-12 |
| Limitations | 19 | Discuss limitations of the study, taking into account sources of potential bias or imprecision. Discuss both direction and magnitude of any potential bias | 13-14 |
| Interpretation | 20 | Give a cautious overall interpretation of results considering objectives, limitations, multiplicity of analyses, results from similar studies, and other relevant evidence | 14 |
| Generalisability | 21 | Discuss the generalisability (external validity) of the study results | 13 |
| **Other information** |  |  |  |
| Funding | 22 | Give the source of funding and the role of the funders for the present study and, if applicable, for the original study on which the present article is based | 15 |

\*Give information separately for exposed and unexposed groups.
**Note:** An Explanation and Elaboration article discusses each checklist item and gives methodological background and published examples of transparent reporting. The STROBE checklist is best used in conjunction with this article (freely available on the Web sites of PLoS Medicine at http://www.plosmedicine.org/, Annals of Internal Medicine at http://www.annals.org/, and Epidemiology at http://www.epidem.com/). Information on the STROBE Initiative is available at http://www.strobe-statement.org.

